# Effectiveness and efficiency of the meningococcal C toddler vaccination and of introducing meningococcal ACWY toddler and adolescent vaccination in Germany

**DOI:** 10.1101/2025.08.01.25332350

**Authors:** Felix Günther, Vanessa Piechotta, Ole Wichmann, Thomas Harder, Alexander Dalpke, Frank G. Sandmann

**Author notes:** **Corresponding author** Felix Günther, Immunization Unit - Team Vaccine/VPD Modelling, Department for Infectious Disease Epidemiology, Robert Koch Institute, Nordufer 20, 13353 Berlin, Germany.

## Abstract

**Background:** In Germany, primary vaccination against invasive meningococcal disease (IMD) in young children is recommended against serotype C (MenC) since 2005 and B (MenB) since 2024. Because of changes in the epidemiology of serogroups C and Y, we re-evaluated the MenC toddler vaccination, also considering scenarios of MenACWY vaccination.

**Methods:** We used a dynamic-transmission model of meningococcal carriage calibrated to national surveillance data for 10-year simulations. We compared MenC vaccination in toddlers aged 12-23 month with scenarios of (i) no MenC/MenACWY vaccination, (ii) MenACWY toddler vaccination, (iii) MenACWY primary adolescent vaccination, and (iv) combined MenACWY toddler plus adolescent booster vaccination. We compared prevented IMD cases, sequelae and deaths. In sensitivity analyses we varied key assumptions like adolescent vaccine uptake.

**Results:** The expected annual mean of 243 (95%-uncertainty interval: 220-258) IMD cases with the MenC toddler vaccination increased by 2.9 (2.2-3.7) cases without MenC vaccination, including 1.2 (0.9-1.5) sequelae and 0.2 (0.2-0.3) deaths. Conversely, IMD cases were reduced by 2.4 (1.7-2.9), 1.7 (0.9-2.6), or 6.8 (5.7-7.6), with MenACWY toddler, primary adolescent, or combined toddler and adolescent vaccination, respectively. Preventing one IMD case required 210,000 (170,000-280,000) MenC or 120,000 (98,000-140,000) MenACWY toddler, 99,000 (84,000-110,000) MenACWY toddler and adolescent, and 72,000 (61,000-86,000) MenACWY primary adolescent vaccinations. Effectiveness of adolescent vaccination increased near-linearly with uptake.

**Conclusions:** Continuing toddler vaccination with MenACWY (or MenC) keeps the IMD burden slightly lower than without toddler vaccination. Yet, introducing MenACWY primary adolescent vaccination was the most efficient scenario, pending sufficient uptake. Serotype carriage requires up-to-date monitoring.

## Introduction

Invasive meningococcal disease (IMD) is a serious condition caused by the gram-negative bacteria *Neisseria meningitidis*, with a case-fatality rate of about 10% [1] and long-term sequelae in up to 20% of cases [2]. Asymptomatic meningococcal infections are widespread in Europe with around 10% in the healthy general population; there is clear variation by age, with carriage prevalence in infants and young children estimated rather low (less than 5%) and peaking at over 20% at around 19 years of age [3, 4]. Nonetheless, IMD remains rare in Germany. Based on national surveillance data the yearly incidence was 0.4 cases per 100,000 individuals between 2012-2015 [1]. After decreases in the years of the coronavirus disease 2019 (COVID-19) pandemic, incidences have returned to pre-pandemic levels in Germany and other countries across Europe [5, 6]. IMD incidence differs by age, with highest incidence among young children and in particular infants; a second peak in incidence exists among adolescents [1, 7]. The strains responsible for the highest disease burden are serogroup B (MenB), followed by serogroups MenY, MenW, and MenC. The relative contributions of the different serogroups change over time due to serotype-specific immunization strategies, disease outbreaks and general epidemiological trends. Similar to other countries, MenY IMD cases have increased in Germany, while the MenC incidence has been decreasing continuously since 2007 [1, 8, 9].

In Germany, routine vaccination against IMD is recommended with a MenC conjugate vaccine in toddlers aged 12-23 month since 2006 [10], and with a protein-based MenB vaccine in infants older than 2 months since 2024 [11]. In 2025, most EU/EEA countries recommend routine infant/toddler vaccination with a mono- or quadrivalent MenC/MenACWY vaccine (18 out of 30), and 11 out of 30 countries also recommend routine adolescent (booster) vaccination [12].

The impact of an additional adolescent MenC or MenACWY booster vaccine at different ages in Germany was investigated before [13, 14], highlighting the need to further investigate the impact of the MenC primary toddler vaccination. In this study, therefore, we examined recent trends in meningococcal epidemiology in Germany to re-evaluate the effectiveness and efficiency of the MenC primary toddler vaccination. We also quantified the potential impact of changing to MenACWY primary toddler vaccination, introducing MenACWY adolescent booster vaccines or switching to MenACWY primary adolescent vaccination. The results were used to inform the national immunization technical advisory group in Germany (Standing Committee on Vaccination, STIKO).

## Methods

### Epidemiological data

IMD is notifiable in Germany and individual case notifications are submitted by local health authorities via the federal state level to the Robert Koch Institute (RKI), the national public health authority for infectious diseases. Aggregated age- and serotype-specific annual case numbers are published by the RKI since 2002 [15]. As IMD is usually severe and requires hospitalisation, no relevant underreporting is expected. To investigate trends in IMD epidemiology, we calculated yearly incidence rates per 100,000 individuals in 3 serogroups (C, AWY; Other/B) from cases reported for the years 2002-2024. These groups are responsible for the highest disease burden in Germany and corresponding to the serogroups targeted by the different available vaccines. We only considered case notifications that met the reference definition [16]. Data were retrieved on April 1, 2025. Missing serotype information was imputed based on the relative serotype distribution in Germany in the respective year.

### Dynamic transmission model

To simulate the effect of different vaccination strategies, we used a dynamic transmission model on the carriage level that was described in detail elsewhere [13]. In short, the model is a time-continuous, deterministic, age- and serogroup-stratified compartmental model. Individuals move between susceptible and serogroup-specific carriage compartments based on an age- and serogroup-specific force of infection (i.e., the rate of acquiring meningococcal carriage of the respective serogroup). Expected yearly IMD cases were derived from multiplying yearly incident new carriers by age and serogroup with case-carrier ratios (CCRs) by age and serogroup. These CCRs were used as scaling factors (in the range 0-1) that correspond to the fraction of incident new carriers that develop IMD. Meningococcal carriage is temporary and individuals returned to the susceptible compartments based on a specified rate (average carriage duration of 6 months). Vaccination was incorporated through additional compartments and can – depending on the vaccine type - reduce the serogroup specific force of infection, as well as the probability of IMD (the CCRs). Vaccine protection is also temporary and waned based on a fixed rate (on average after 4 years for individuals <12 years of age, and 10 years for >=12 years age). Vaccinated but unprotected individuals were subject to the same rate of carriage acquisition and probability of IMD as unvaccinated individuals. Ageing and vaccination were implemented through time-discrete, yearly step-changes. The model was calibrated to German IMD case counts by year and age from 2005-2019 via negative binomial maximum likelihood estimation. Parameter uncertainty from the fitting was propagated through the forward-simulation of vaccination scenarios. For more details and assumptions, see Supplementary table M1 and [13].

### Simulated vaccination scenarios

We simulated five different vaccination scenarios over a 10-year simulation period:

- Scenario 1: MenC primary toddler vaccination in the second year of life (month 12-23)
- Scenario 2: No further MenC/MenACWY vaccination at any age
- Scenario 3: MenACWY primary toddler vaccination in the second year of life (month 12-23)
- Scenario 4: MenACWY primary toddler vaccination in the second year of life (month 12-23) plus MenACWY booster vaccination at age 16
- Scenario 5: MenACWY primary adolescent vaccination at age 16 and discontinuation of MenC/MenACWY toddler vaccination

Scenario 1 matched the current recommendation in Germany (base case). To compare the efficiency of the MenC primary toddler vaccination, we also simulated a scenario without MenC/MenACWY vaccination in future years (scenario 2). The scenarios of primary toddler vaccination assumed a vaccine uptake of 80% as observed at the end of the second year of life in 2019-2021 [17]. For adolescent vaccinations (scenarios 4 and 5) we assumed an uptake of 40%, similar to the uptake for the adolescent human papillomavirus (HPV) vaccination in Germany [18]. The age of 16 years was chosen here based on results of a previous study [13]. In all scenarios we accounted for the newly introduced MenB toddler vaccination in Germany by downscaling expected IMD cases among infants and young children assuming 80% vaccine uptake, a vaccine effectiveness (VE) against MenB/Other IMD of 75% and an average duration of protection of 10 years.

### Modelled outcomes

We quantified results by age and serogroup in terms of the number of expected IMD cases, IMD cases with long-term sequelae and deaths. Probabilities for 16 different sequelae were sourced from the published literature [19, 20]. For deaths we multiplied cases with the estimated case-fatality ratios (CFRs) by age and serogroup in Germany [13].

Afterwards, we estimated the associated numbers-needed-to-vaccinate (NNVs) to prevent one case with or without sequelae or death by comparing the results from alternative vaccination strategies with the results of the current recommendation (MenC primary toddler vaccination). To compare the efficiency of the current recommendation and other vaccination strategies, we also compared results to the scenario without any vaccination (scenario 2), and afterwards in a fully incremental analysis (i.e., against the next-most efficient strategy).

### Sensitivity analyses

To investigate key modelling assumptions, we first restricted results to serogroups MenC and MenAWY and omitted serogroup Other/B, which corresponds to an assumption of no serogroup replacement when changing the MenC/MenAWY vaccination strategy. Second, we extended the simulation period from 10 to 30 years. Third, we used a static model based on reported IMD incidences from the years 2023-2024 that does not depend on pre-pandemic trends but a constant serogroup and age-specific incidence as observed post-COVID-19-pandemic. This approach does not include indirect effects (for more details see Supplementary text M1). In this model, we also varied the age of adolescent vaccination.

## Results

### Epidemiological trends

According to notification data, the yearly IMD incidence decreased from the pre-vaccination period (2002-2004) to the pre-COVID-19 pandemic years (2017-2019) from 0.84 to 0.34 cases per 100,000 individuals. During the initial three years of the COVID-19 pandemic (2020-2022), the incidence was substantially reduced (0.14 per 100,000), before it returned to pre-COVID-19 pandemic levels (0.34 per 100,000, 2023-2024; Fig 1A).

**Figure 1:**
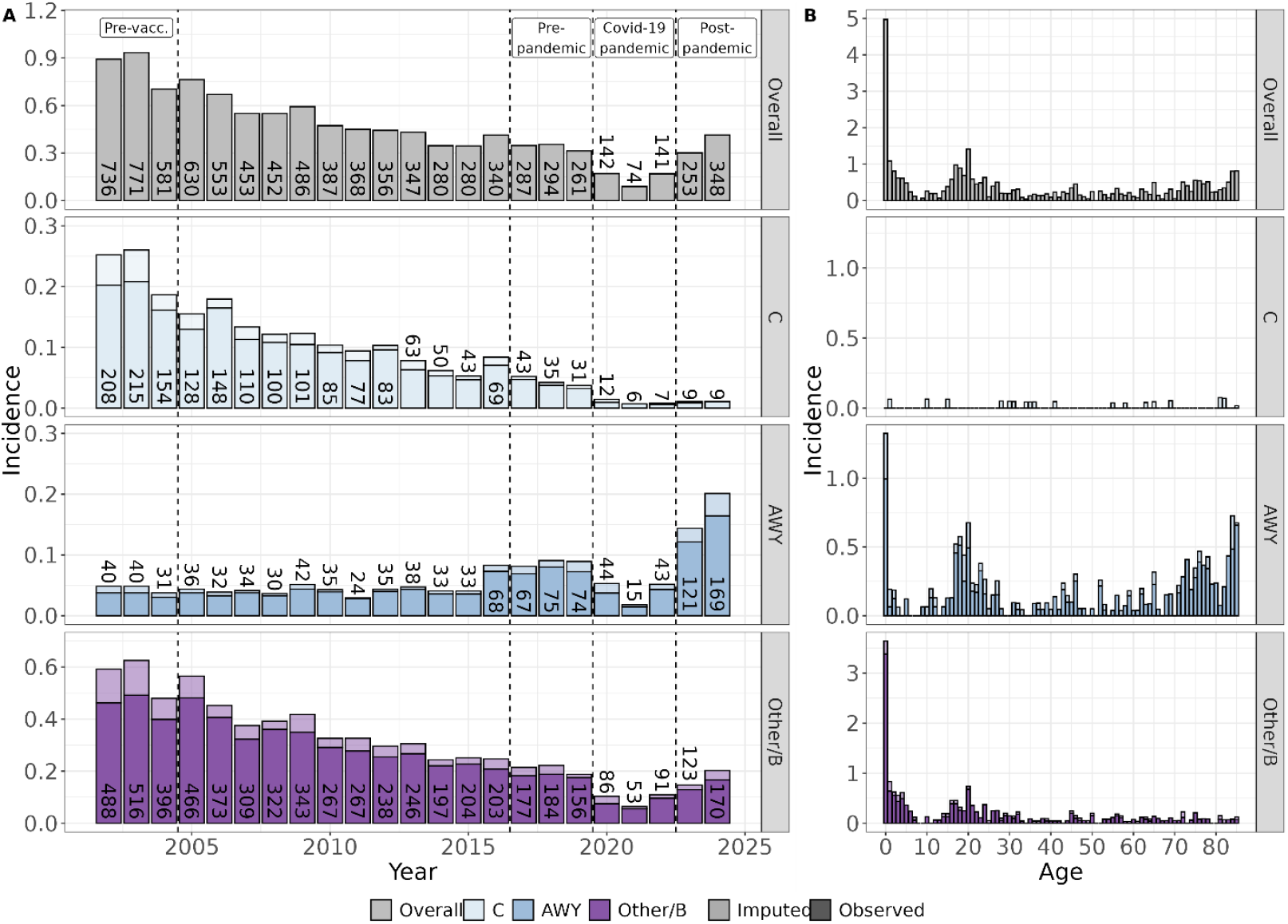
Reported IMD incidence per 100,000 individuals in Germany by year, from 2002-2024 (panel A) and by age in the post-Covid-19 pandemic years 2023-2024 (panel B). IMD: invasive meningococcal disease. Bars represent the reported IMD incidence per 100,000 person-years overall and separate in three serogroups (MenC, MenAWY, and Other/B). Panel A shows overall yearly incidence from national surveillance in 2002-2024; panel B shows the total IMD incidence for one-year age groups 0-84, and 85+ years in 2023-2024. Dotted lines in panel A mark specific time periods mentioned in the text, including the main period of the COVID-19 pandemic (2020-2022). Absolute numbers correspond to reported case counts. Cases with missing serogroup information where imputed based on the relative serogroup-distribution in the respective year and visualised in a lighter colour. Note the different y-axis range in the sub-panels.

During 2002-2024, 67% of all IMD cases were due to serogroup Other/B, 20% due to MenC and 13% due to MenAWY (Fig 1A). However, the IMD incidence decreased substantially for MenC and Other/B over 2002-2019, while it approximately doubled for MenAWY until 2019. After a reduction across serogroups during the pandemic, IMD numbers remained very low for MenC in 2023-2024, while they returned to pre-pandemic levels for Other/B and increased substantially for MenAWY. In 2023-2024, the incidence of MenAWY was nearly as high as for Other/B (0.173 and 0.174 per 100,000 individuals), and much higher than MenC (0.01 per 100,000) (Fig 1A).

The IMD incidence was highest among infants, with a second peak in ages 16-18 years (Fig 1B, SuppFig 1). This age-pattern of IMD incidence was relatively consistent across serogroups. MenAWY incidence also increased among older adults and reached a third peak above age 70. In children aged 1-5 years, which includes the target age of the MenC primary toddler vaccination, MenC incidence decreased to near-zero until 2018 (SuppFig 2). In the age- and serogroups targeted by a MenACWY toddler or adolescent vaccination, post-pandemic incidence was highest in adolescents due to MenAWY IMD (Figure 1B). Overall, pre-pandemic incidence trends in age- and serogroups continued after the interruption of the pandemic (SuppFig2).

### Effectiveness of vaccination strategies

During the 10-year model simulation period, the average number of expected yearly IMD cases was 243 (220-258) for the baseline scenario of continuing MenC primary toddler vaccination, i.e. 0.29 (0.26-0.31) cases per 100,000 individuals (Fig 2A). The serogroup-specific epidemiological trends were continued during the simulation period (Fig. 2C). Simulated yearly incidence per 100,000 individuals was highest for the Other/B serogroup with 0.18 (0.14 – 0.20), followed by MenAWY with 0.08 (0.07-0.09) and MenC with 0.03 (0.03-0.04). Towards the end of the 10-year period, expected IMD counts for MenAWY were almost as high as for serogroup Other/B (Fig 2C).

**Figure 2:**
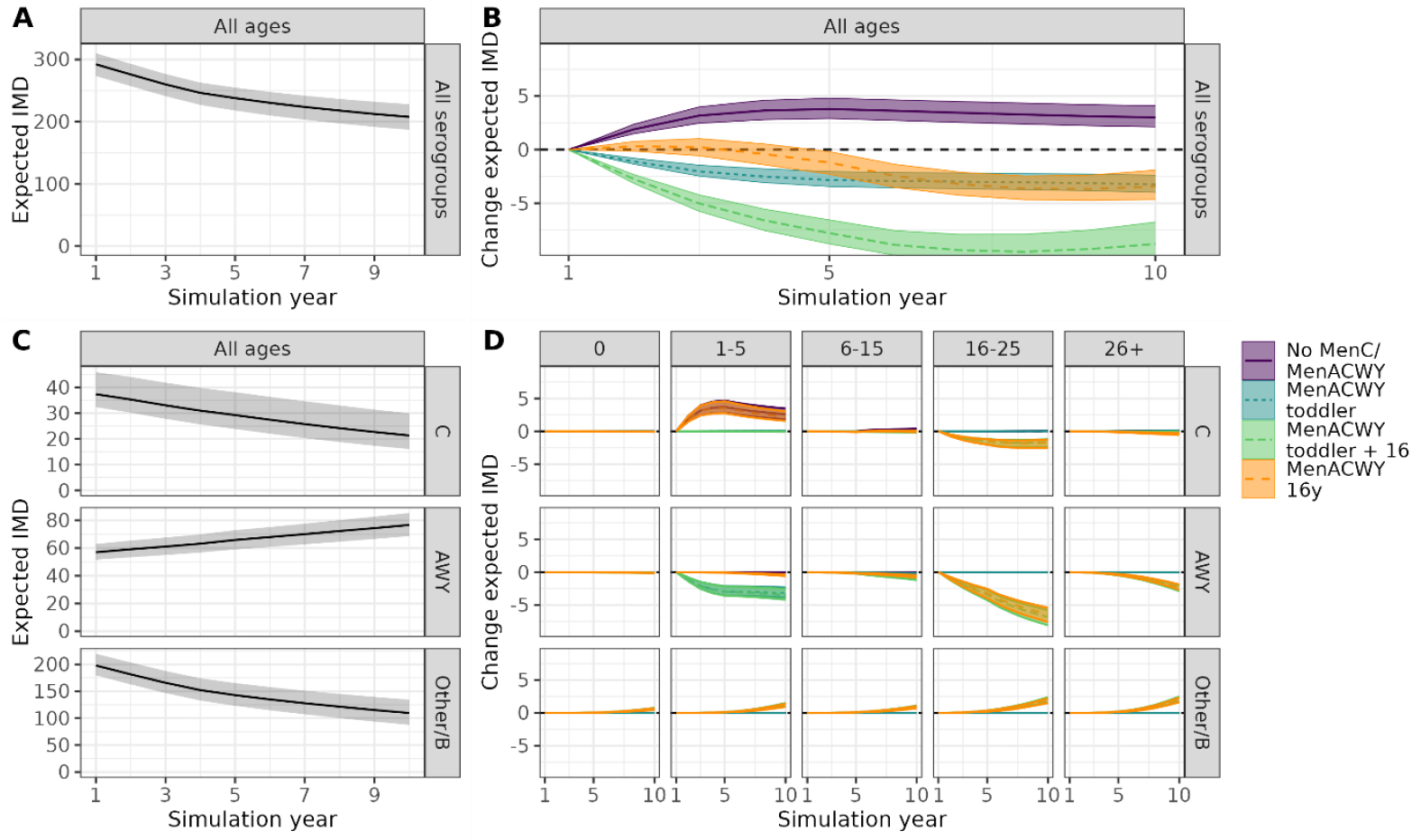
Simulated expected number of annual IMD cases in Germany over 10-years when continuing MenC toddler vaccination (panel A and panel C), and the expected changes in annual IMD cases without the MenC/MenACWY infant vaccination and when switching to the different MenACWY vaccination strategies (panel B and panel D). IMD: invasive meningococcal disease. Panel A shows the expected yearly IMD case counts during the 10-year simulation period for the baseline simulation scenario of continuing MenC toddler vaccination. Panel B shows the difference in expected case counts for four alternative vaccination strategies (color-coded). Panel C shows extrapolated case counts for MenC toddler vaccination separated into three serogroups. Panel D shows changes in expected case counts due to different vaccination strategies in 5 age and 3 serogroups. Shown are model-based expected case counts and 95%-uncertainty intervals.

As compared to the scenario without any MenC/MenACWY vaccination (scenario 2), continuing the MenC primary toddler vaccination (scenario 1) prevented an estimated mean of 2.9 (2.2-3.7) cases, 1.2 (0.9-1.5) sequelae and 0.2 (0.2-0.3) deaths annually (Fig 2B, Table 1).

**Table 1:** Effectiveness and efficiency of the different vaccination strategies (annual mean values over the 10-year time horizon + 95% uncertainty intervals).

| A) Comparison against MenC toddler |  |  |  |  |  |  |  |
| --- | --- | --- | --- | --- | --- | --- | --- |
|  | Prevented cases |  |  | NNV |  |  |  |
|  | IMD | Sequelae | Death | Diff. Vacc. | IMD | Sequelae | Death |
| MenACWY toddler | 2.4<br>(1.7;2.9) | 0.9<br>(0.7;1.1) | 0.1<br>(0.1;0.1) | 0 | _* | _* | _* |
| MenACWY toddler + 16y | 6.8<br>(5.7;7.6) | 2.7<br>(2.3;3.0) | 0.5<br>(0.5;0.6) | 334,000 | 49,000<br>(44,000;59,000) | 120,000<br>(110,000;150,000) | 610,000<br>(550,000;720,000) |
| MenACWY 16y | 1.7<br>(0.9;2.6) | 0.7<br>(0.4;1.1) | 0.2<br>(0.1;0.3) | -284,000 | _* | _* | _* |
| B) Comparison against no MenC/MenACWY vaccination |  |  |  |  |  |  |  |
|  | Prevented cases |  |  | NNV |  |  |  |
|  | IMD | Sequelae | Death | Diff. Vacc. | IMD | Sequelae | Death |
| MenC toddler | 2.9<br>(2.2;3.7) | 1.2<br>(0.9;1.5) | 0.2<br>(0.2;0.3) | 618,000 | 210,000<br>(170,000;280,000) | 530,000<br>(410,000;700,000) | 2,500,000<br>(2,000,000;3,400,000) |
| MenACWY toddler | 5.3<br>(4.4;6.3) | 2.1<br>(1.7;2.5) | 0.4<br>(0.3;0.4) | 618,000 | 120,000<br>(98,000;140,000) | 300,000<br>(250,000;360,000) | 1,700,000<br>(1,400,000;2,100,000) |
| MenACWY toddler + 16y | 9.7<br>(8.5;11.3) | 3.9<br>(3.4;4.5) | 0.8<br>(0.7;0.9) | 951,000 | 98,000<br>(84,000;110,000) | 240,000<br>(210,000;280,000) | 1,200,000<br>(1,000,000;1,400,000) |
| MenACWY 16y | 4.6<br>(3.9;5.4) | 1.9<br>(1.6;2.2) | 0.5<br>(0.4;0.5) | 334,000 | 72,000<br>(61,000;86,000) | 170,000<br>(150,000;210,000) | 730,000<br>(620,000;860,000) |
| C) Comparison against MenACWY 16y |  |  |  |  |  |  |  |
|  | Prevented cases |  |  | NNV |  |  |  |
|  | IMD | Sequelae | Death | Diff. Vacc. | IMD | Sequelae | Death |
| MenC toddler | -1.7<br>(-2.6;-0.9) | -0.7<br>(-1.1;-0.4) | -0.2<br>(-0.3;-0.1) | 284,000 | -** | -** | -** |
| MenACWY toddler | 0.6<br>(-0.3;1.7) | 0.2<br>(-0.2;0.6) | -0.1<br>(-0.2;0) | 284,000 | 450,000<br>(160,000; - **) | 1,800,000<br>(470,000; - **) | -** |
| MenACWY toddler + 16y | 5.1<br>(4.2;6) | 2<br>(1.6;2.3) | 0.3<br>(0.3;0.4) | 618,000 | 120,000<br>(100,000;150,000) | 310,000<br>(260,000;380,000) | 1,900,000<br>(1,500,000;2,200,000) |

IMD: invasive meningococcal disease, NNV: number needed to vaccinate. Results from the main analysis are aggregated over all sergroups and show the net-effect of the vaccination strategies, also accounting for serotype replacement. For results without serotype replacement, see Supplementary Table 1.
\*We do not report NNVs in case of reduced disease burden from the same or a reduced number of vaccinations
\*\* No NNV is reported in case of increased disease burden and an increased number of vaccinations

Switching from MenC (scenario 1) to MenACWY primary toddler vaccination (scenario 3) prevented on average 2.4 (1.7-2.9) additional IMD cases per year (Fig 2B, Table 1). The reduction was mostly due to the direct effects of reduced MenAWY cases (Fig 2D, 1-5 years of age).

Switching from MenC (scenario 1) to MenACWY primary toddler vaccination and a MenACWY adolescent booster (scenario 4) was most effective and prevented on average 6.8 (5.7-7.6) additional IMD cases per year (Fig 2B, Table 1). Next to the direct effects in MenC and MenAWY (Fig 2D, 16-25 years of age), indirect effects also slightly reduced IMD cases of AWY in other ages. However, serogroup-replacement effects occurred towards the end of the 10-year simulation (Fig 2D; cf. serogroup Other/B).

Switching from MenC primary toddler vaccination (scenario 1) to MenACWY primary adolescent vaccination (scenario 5) reduced expected IMD counts by an average of 1.7 (0.9-2.6) cases per year (Fig 2B, Table 1). A small increase in expected MenC IMD cases in young children was seen, in particular during the first years of the simulation period, and an increasing reduction in expected MenC and MenACWY cases in adolescents (Fig. 2D). The effects of MenACWY primary adolescent vaccination or adolescent booster vaccination were comparable for the adolescents and adults, and the indirect effects were very similar (Fig. 2D).

### Efficiency of vaccination strategies

Switching the primary toddler vaccination from MenC (scenario 1) to MenACWY (scenario 3) reduced the disease burden for the same number of vaccinations, making the MenACWY vaccination more efficient. MenACWY primary toddler vaccination plus adolescent booster (scenario 4) resulted on average in 334,000 additional vaccinations per year (assuming a 40% booster uptake and compared to scenario 1). Compared to MenC primary toddler vaccination (scenario 1), the number of vaccinations to prevent one IMD case was 49,000 (44,000-59,000), 120,000 (110,000-150,000) per sequelae and 610,000 (550,000-720,000) per death prevented (Table 1A). A switch to MenACWY primary adolescent vaccination (scenario 5) prevented a larger disease burden than the MenC primary toddler vaccination (scenario 1) at 284,000 fewer vaccinations per year (due to a lower uptake assumed), and would therefore be more efficient.

As compared to the hypothetical scenario of no MenC/MenACWY vaccination (scenario 2), the most efficient strategy was MenACWY primary adolescent vaccination (scenario 5) with an NNV to prevent one IMD case of around 72,000 (61,000-86,000), followed by MenACWY toddler vaccination plus adolescent booster (scenario 4) with an overall NNV to prevent one IMD case of around 98,000 (84,000-110,000) (Table 1B, Fig 3). Incrementally, moving from an adolescent MenACWY primary vaccination (scenario 5) to MenACWY toddler vaccination plus adolescent booster (scenario 4) the NNV to prevent one IMD case was 120,000 (100,000-150,000) (Table 1C).

**Figure 3:**
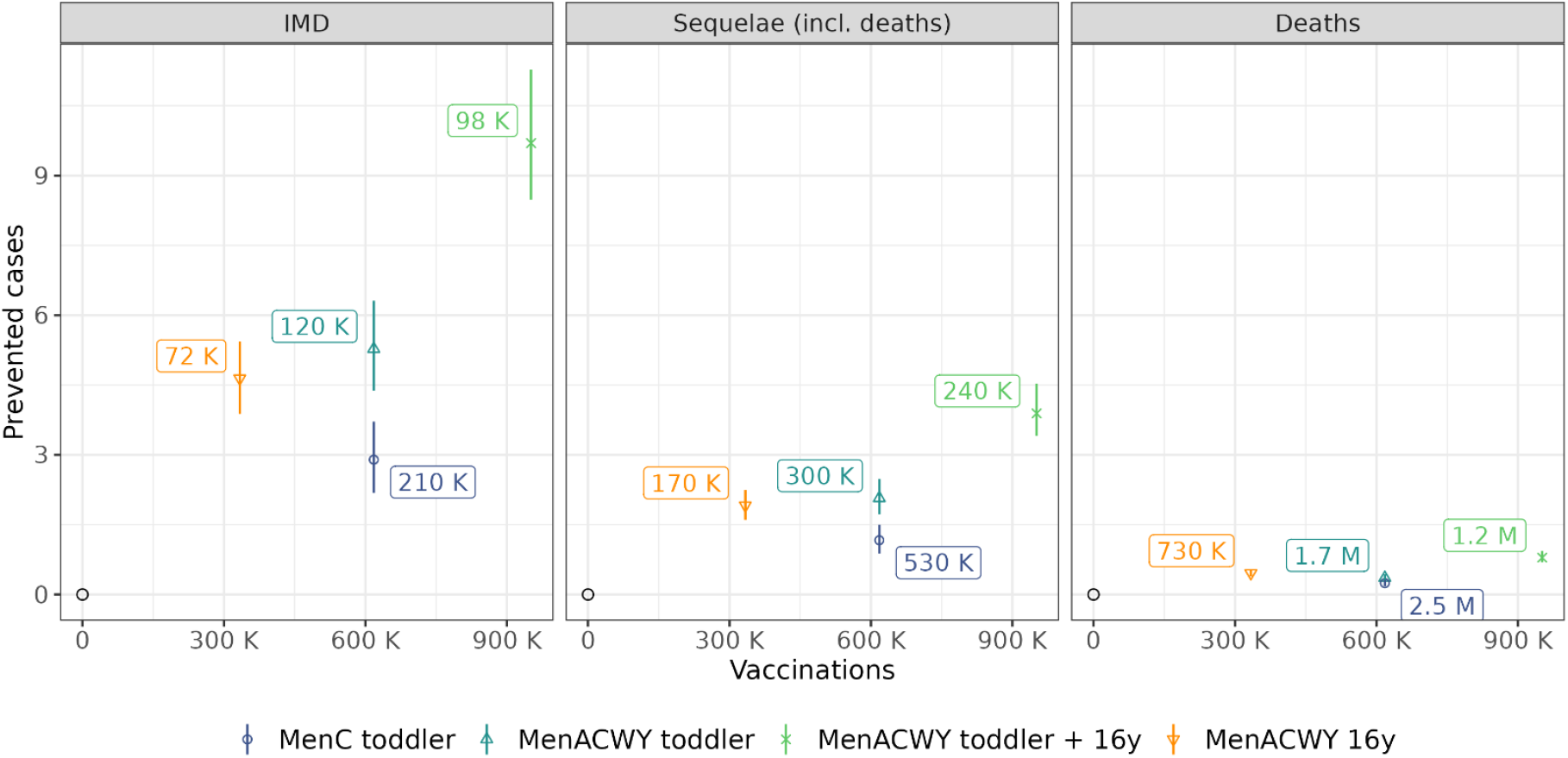
Effectiveness and efficiency of the different vaccination strategies in terms of the number of vaccinations, the number of prevented cases, and the number needed to vaccination to prevent one case (IMD, sequelae, deaths). IMD: invasive meningococcal disease, K: kilo (i.e., in thousands), M: in millions, UI: uncertainty interval. Shown are average annual vaccination numbers and prevented expected cases (yearly, point estimates + 95%UIs) for four different vaccination strategies compared to the no MenC/MenACWY vaccination strategy over the 10-year simulation period. Numbers in labels represent the respective number needed to vaccinate to prevent one case (point estimate).

### Results of the sensitivity analyses

The uptake of the adolescent ACWY vaccination directly impacted the expected yearly number of prevented IMD cases, which changed (near-)linearly with uptake. MenACWY primary adolescent vaccination would need to achieve an uptake of about an estimated 46% to prevent a similar number of IMD cases than the switch to MenACWY primary toddler vaccination. The efficiency in terms of the NNV remained relatively constant (Figure 4, panel “Main”).

**Figure 4:**
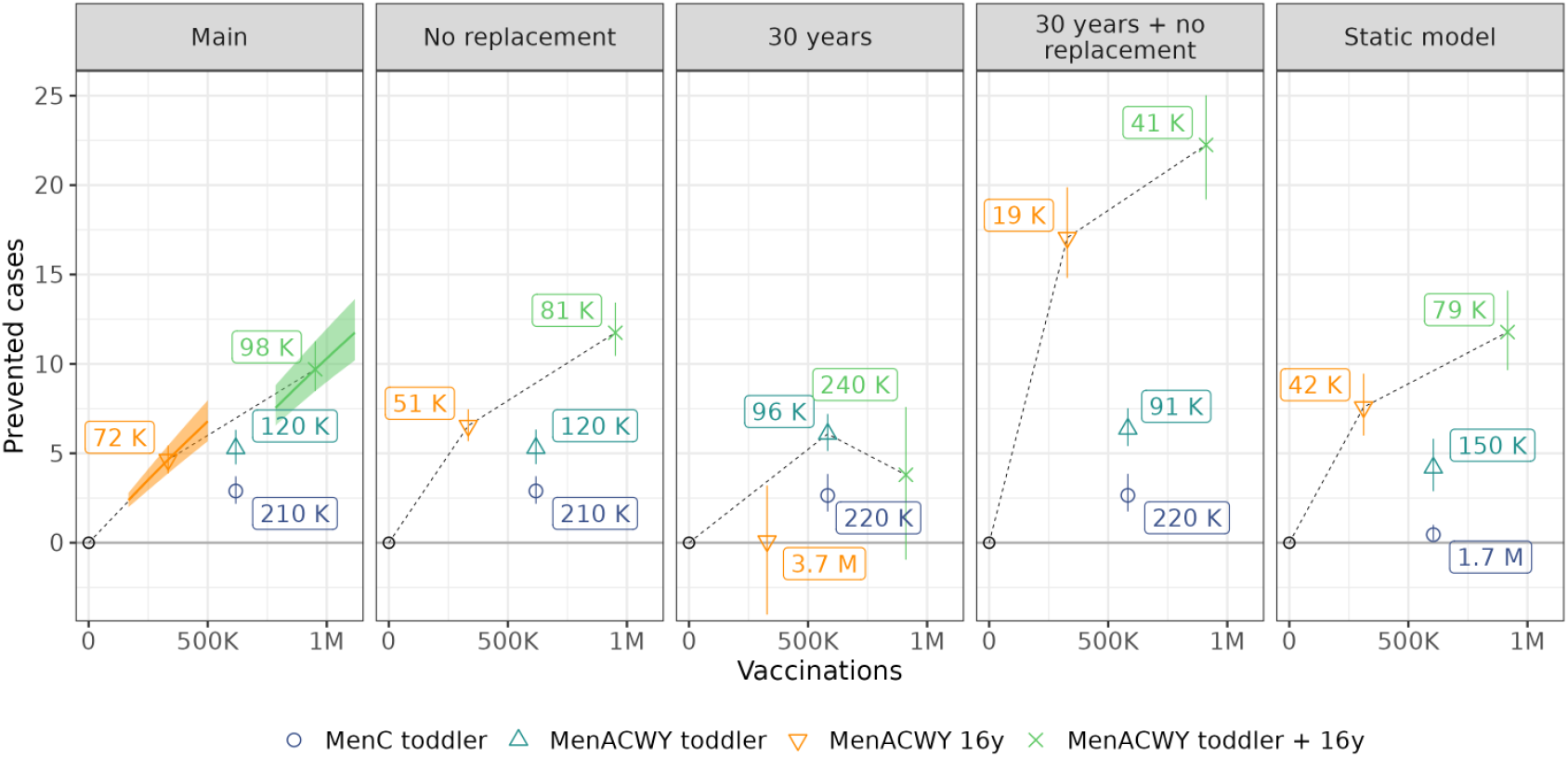
Sensitivity analysis of the effectiveness and efficiency of the different vaccination strategies in terms of the number of vaccinations, the number of prevented cases, and the number needed to vaccination to prevent one IMD case. IMD: invasive meningococcal disease, K: kilo (i.e., in thousands), M: in millions, UI: uncertainty interval. Shown are average annual vaccination numbers and prevented expected IMD cases (yearly, point estimates + 95%UIs) for four different vaccination strategies compared to the no MenC/MenACWY vaccination strategy. Each panel shows different assumptions for the simulation of vaccination strategies: Panel “Main” shows the results from the main analysis, corresponding to Figure 3, Panel “IMD”. The orange/green lines and ribbons show simulation results when varying the MenACWY vaccine uptake among adolescents from 20% to 60%. Panel “No replacement” corresponds to the main analysis ignoring expected IMD cases in the Other/B serogroup, this corresponds to an assumption of no serotype replacement towards other serogroups. Panel “30 years” shows simulation results for a 30-year period, “30 years + no replacement” are results in the longer simulation period without serotype replacement towards serogroup Other/B. Panel “Static model” shows results for a static model of vaccine effects assuming constant incidence based on post-pandemic incidence data in Germany from 2023-2024. Numbers in labels represent the number needed to vaccination to prevent one case as compared to no MenC/MenACWY vaccination (point estimates).

The effectiveness and efficiency of the MenACWY adolescent vaccination was reduced by serotype replacement. Without serotype replacement, adolescent primary vaccination would prevent a mean of 6.5 (5.7, 7.5) IMD cases per year, and the NNV to prevent one IMD case would be approximately 29% lower at 51,000 (45,000, 59,000). Strain replacement did not play a substantial role for toddler vaccination in the simulations due to low carriage prevalence in this age group (Supp. Table 1A, Fig 4).

When simulating the effects of different vaccination strategies over a 30-instead of a 10-year period, the influence of indirect effects became larger (SuppFig 3). The simulation period had little impact for toddler vaccination (SuppTab 1). For the MenACWY adolescent vaccination, due to serotype replacement, the positive effect of prevented IMD cases for MenACWY was largely offset by an increase in expected IMD cases in Other/B. When assuming no serotype replacement, the indirect protection from MenACWY adolescent vaccination accumulated over time, leading to a higher effectiveness and efficiency over the 30-year simulation period, e.g., resulting in an average yearly number of prevented IMD cases of 17.1 (14.8, 19.9) and a NNV to prevent one IMD case of 19,000 (16,000, 22,000) compared to no MenC/MenACWY vaccination (SuppTab 1B, Fig 4).

Based on a static model assuming a time-constant serogroup-specific incidence as observed in 2023-2024, the current MenC toddler vaccination strategy was less effective and efficient due to the very low, near-zero MenC incidence among young children. Conversely, the vaccination strategies that included adolescent MenACWY vaccination were more effective and efficient due to slightly higher incidences and no serotype replacement (Figure 4). Adolescent MenACWY vaccination was most efficient at age 17 (SuppFig 4).

## Discussion

In this study, we evaluated the effectiveness and efficiency of continuing the current MenC primary toddler vaccination in Germany and modelled the impact of alternative MenACWY vaccination strategies. Our results showed that all vaccination strategies were effective, with the current MenC primary toddler vaccination estimated to prevent a mean of three IMD cases annually in Germany, and one death every five years. Switching from MenC to MenACWY primary toddler vaccination can prevent additional IMD cases, in particular given the decreasing trend in MenC and stable or increasing trend in MenW and MenY incidence. The introduction of a MenACWY adolescent booster alongside a MenACWY primary toddler vaccination was the most effective strategy preventing the largest absolute burden. However, MenACWY primary adolescent vaccination was the most efficient strategy, saving on hundred-thousands of vaccine doses per year.

Our results can be explained largely due to the relatively high IMD incidence among adolescents and the very low MenC incidence, and the potential for indirect effects through reducing meningococcal carriage in the high-prevalence group of adolescents. However, the simulations also suggested relevant serotype replacement, where the positive effects of prevented IMD cases for MenACWY were (partly) offset by increased IMD cases of other serogroups. These effects were more pronounced for longer simulation periods, and can reduce the effectiveness and efficiency of MenACWY adolescent vaccination substantially. Nonetheless, our most optimistic NNV of an estimated 19,000 MenACWY primary adolescent vaccinations to prevent one IMD case over 30 years and without serotype replacement compares to a similar NNV of about 12,000 for the MenB infant vaccination (which could be slightly higher by now, too, given the decreasing trend for MenB) [11]. Overall, our model results were robust to other changes in key assumptions.

Compared to vaccinations against other diseases, the estimated efficiency of meningococcal vaccination, particularly the MenC toddler vaccination, is relatively small due to the low incidences. Similar incidence patterns are reported across EU/EEA Member States, with only 55 confirmed IMD cases in 2022 being caused by MenC (corresponding to 6%, and no more than 20% in the unlikely situation of attributing all cases with unknown serogroup to MenC). In infants and toddlers, serogroup B caused nearly 80% of confirmed IMD cases, followed by serogroup W and only in third place serogroup C (at less than 5%) [5].

Few routine vaccinations are recommended for adolescents in Germany [21]. Approximately 55% of 15-year-old girls and 34% of 15-year-old boys in Germany have been vaccinated against HPV with two doses [18]. Due to this and limited experience with routine vaccination among adolescents, it is difficult to anticipate how high the uptake of a MenACWY adolescent vaccination would be. An uptake of 40%, as in our main analysis, appears to be a realistic and rather conservative assumption, with an uptake of about 46% required to be on par with the effectiveness of switching to MenACWY primary infant vaccination. Based on our model, the effectiveness of adolescent MenACWY vaccination increases in a range of uptake (20-60%) near-linearly. In practice, spatial or social differences can influence the overall impact of vaccination.

In response to increasing MenW IMD cases before the SARS-CoV-2 pandemic, several countries have recommended MenACWY vaccination, also among adolescents in England, the Netherlands, and Australia. Recently, the UK switched to recommending primary MenACWY immunization among adolescents, instead of infants/toddlers [22, 23]. A modelling study supported the decision to discontinue the MenC primary infant vaccination in the UK, reaching similar conclusions for the UK as our study for Germany. In addition, they also showed the marginal benefit from administering MenACWY at 3-months instead 12-months of age in preventing an estimated 8 out of 11 MenACWY cases in the first year of life (72.7%) [24]. In practice switching to MenACWY at 3-months would also require a change in the number of doses (2+1 vs. 1 dose scheme). Results from post-implementation surveillance generally indicate high effectiveness in the target groups and also other age-groups [25-27]. Recent post-pandemic data from England also indicate continued protection from the adolescent MenACWY vaccination in serogroup MenACWY [28].

### Strengths and limitations

We used a dynamic transmission model that is first to describe introducing primary adolescent MenACWY vaccination in Germany, which was also used to inform deliberations of the NITAG. Our study was based among others on the most recent national surveillance data for Germany, and we explored uncertainty in comprehensive sensitivity analyses.

Up-to-date data on meningococcal carriage remain scarce. In the UK, low MenCWY carriage prevalence was reported in adolescents after introduction of MenACWY adolescent vaccination [29]. There is currently no epidemiological evidence of serogroup replacement following MenACWY vaccination for adolescents. However, available data is limited and it is difficult to disentangle the effects of different factors on serogroup-specific disease burden. In our model, replacement effects started to appear only several years after introducing adolescent vaccination. Previously, it was shown how the magnitude of serogroup replacement depends on the (relative) carriage prevalence of different meningococcal serogroups, case-carrier ratios and the vaccine effectiveness against carriage [13]. Data on these drivers are limited, with no recent data for Germany on age- and serogroup-specific meningococcal carriage prevalence. For our simulations, we extrapolated serogroup-specific trends from the period before the COVID-19 pandemic, and the first post-pandemic data from 2023 and 2024 show that this is plausible, even if reported MenAWY cases are slightly higher than expected in the extrapolation. Our estimates of the effectiveness and efficiency of quadrivalent MenACWY vaccination may therefore be conservative - however, future developments are unknown, including the novel emergence and resurgence of serogroups from importations into Germany, and may further influence effectiveness and efficiency. Overall, the modelling clearly indicates the need for detailed monitoring of epidemiological developments and more specific studies on, e.g., the carriage prevalence. As serogroup replacement due to indirect effects of the vaccination was an important aspect for vaccine efficiency, continuous serotype surveillance systems must be in place.

In conclusion, switching from MenC to MenACWY primary toddler vaccination is estimated to be more effective. Introducing an additional MenACWY adolescent booster vaccine is the most effective strategy, while moving to MenACWY primary adolescent vaccination is most efficient – if sufficient levels of uptake in adolescents are achieved. Moreover, a switch from infant to adolescent primary MenACWY immunization would free up a slot in the crowded immunization schedule among young children that could improve uptake and help closing gaps for other vaccines in this age-group.

## Acknowledgments section

**Acknowledgements**

The authors thank the members of the meningococcal working group at the German National Immunization Technical Advisory Group called ‘Standing Committee on Vaccination’ (STIKO) for helpful comments.

## Author contributions (based on CRediT author statement)

Felix Günther: Conceptualization, Methodology, Software, Validation, Formal analysis, Investigation, Data Curation, Writing - Original Draft, Writing - Review & Editing, Visualization

Vanessa Piechotta: Conceptualization, Methodology, Investigation, Writing - Review & Editing

Ole Wichmann: Conceptualization, Methodology, Writing - Review & Editing, Funding acquisition

Thomas Harder: Conceptualization, Methodology, Writing - Review & Editing, Funding acquisition

Alexander Dalpke: Conceptualization, Methodology, Writing - Review & Editing

Frank G. Sandmann: Conceptualization, Methodology, Validation, Formal analysis, Writing - Original Draft, Writing - Review & Editing, Supervision, Project administration

## Data availability

Code and data for reproducing the results of the modelling study will be made available in a public repository upon final publication of the manuscript. Meningococcal IMD case numbers are available from Survstat@RKI (https://survstat.rki.de/).

