## Supplemental Material for "Effectiveness and efficiency of the meningococcal C toddler vaccination and of introducing meningococcal ACWY toddler and adolescent vaccination in Germany"

### Supplementary material: Methods

#### Supplementary table M1. Assumptions and data sources used in the dynamic transmission model

| Type | Description/Value | Reference(s)/Comments |
|---|---|---|
| IMD case numbers used for calibration | IMD case numbers by age and serogroup from years 2005-2019, derived from national surveillance data (Survstat@RKI) | Survstat@RKI |
| Vaccination numbers used for calibration | Vaccination numbers collected at RKI (KV-Impfsurveillance) | [1] |
| Case-carrier ratio (CCR) | Age- and serogroup-specific CCR derived from cross-sectional carriage data and IMD incidence in Germany. | Carriage data:<br>[2-4]<br>IMD incidence 2002-2005:<br>Survstat@RKI<br>Details on derivation and operationalization in [5] |
| Mean duration of carriage | 6 months | Sensitivity analysis in [5] showed little impact |
| Force of infection (FOI) | Age- and serogroup-specific FOI estimated during model calibration. | Details in [5] |
| Vaccine efficacy | Parameters from fitting an exponential waning model to data on real-world vaccine efficacy:<br>- Protection against carriage (among vaccinated and protected):<br>MenC: 0.85/0.0/0.0 vs. MenC/AWY/Other<br>MenACWY: 0.16/0.16/0.0 vs. MenC/AWY/Other<br>- Protection against IMD among (incident) carriers:<br>MenC: 1.0/0.0/0.0 vs. MenC/AWY/Other<br>MenACWY: 1.0/1.0/0.0 vs. MenC/AWY/Other<br>- Duration of vaccine-induced protection:<br>MenC and MenACWY: 4 years up to age 12, 10 years afterwards<br>- Corresponding total protection against MenC or MenACWY IMD (age <12 years):<br>88%, 69%, 54%, 38% (during year 1, 2, 3, and 4-5 after vaccination)<br>- Corresponding protection against carriage in first year after vaccination:<br>MenC: 75.2%, MenACWY: 14.2% | Real-world VE estimates from post licensure surveillance after introducing routine infant immunization program in UK: [6]<br><br>Details and further references in [5] |
| Demographic data | Official population data by Destatis up until 2022 and population projections for simulations up until 2049 | [7, 8] |
| Contact frequencies | Polymod | [9] |
| Sequelae and fatalities among IMD cases | Published estimates of the probabilities of 16 different long-term health outcomes for sequelae and national surveillance data from 2002-2019 (Survstat@RKI) for fatalities. | [10]<br>Survstat@RKI<br>Details in [5] |

**Supplementary text M1. Static model used in the sensitivity analysis**

To simulate the effects of alternative vaccination strategies based on a static model in sensitivity analysis and to compare with the dynamic-transmission model (see Methods section of the main text), we firstly estimated a smoothed incidence of IMD by age and serogroup using a generalized additive model (GAM) based on reported IMD case counts from 2015-2024. This time period covered three years before the SARS-CoV-2 pandemic (2015-2019), the three main years of the pandemic (2020-2022), and the most recent (complete) post-pandemic years (2023-2024). We estimated a negative binomial GAM with the yearly number of IMD cases by age (in years) and serogroup as outcome, and log of the corresponding population size as offset (to estimate the incidence rate). We included a smooth age-effect by splines separately for the pre-pandemic (2015-2019), pandemic (2020-2022) and post-pandemic (2023-2024) period. The estimated incidence rate by age is illustrated in Supplementary Figure M1.

To estimate the direct effects of different vaccination strategies with the static model, we derived age-specific fractions of individuals protected against serogroup-specific IMD from vaccination for the different strategies. In line with the dynamic transmission model, we assumed exponential waning of immunity, and derived the population level protection against IMD of serogroup $MenX$ with $X \in \{C, AWY, Other/B\}$ in age-group $a \in \{0, 1, \dots, 85\}$ from vaccination strategy $i$ as

$$p_i^{MenX}(a) = \int_{a}^{a+1} I(t > vacc\_age_i) e^{-r_i \times (t - vacc\_age_i)} \times upt_i \times ve_i^{MenX} dt$$

Where $vacc\_age_i$ is the age of vaccination in strategy $i$ (in years), $upt_i$ the assumed uptake in strategy $i$ (as proportion), $ve_i^{MenX}$ is the vaccine effectiveness against IMD of serogroup $MenX$ for the vaccine used in strategy $i$ (as proportion) and $r_i$ waning of protection for the vaccine used in strategy $i$ (as rate in years). Parameters of this model for the different vaccination strategies are given in Supplementary table M2. For the strategy combining infant and adolescent MenACWY vaccination the fraction of individuals protected of the infant and adolescent vaccination strategies are added, which is reasonable for the static model given the different target (age) groups of both strategies and the focus on direct effects of vaccination.

To estimate effects of strategies we used in a first step the model-based, expected post-pandemic incidence in age-group $a$ and serogroup $MenX$ (which corresponds the incidence under the status quo scenario MenC infant vaccination) $inc_1^{MenX}(a)$, and derived an expected incidence under a hypothetical "no vaccination" scenario based on rescaling with the fraction of protected from

$$inc_0^{MenX}(a) = \frac{inc_1^{MenX}(a)}{(1 - p_1^{MenX}(a))}$$

The expected incidence of other vaccination strategies $i$ are subsequently derived from

$$inc_i^{MenX}(a) = \left(1 - p_i^{MenX}(a)\right) \times inc_0^{MenX}(a)$$

The overall (yearly) number of prevented cases is then derived from summing the difference of expected IMD cases of two vaccination strategies over all age- and serogroups.

*Supplementary figure M1: IMD incidence in Germany by age and serogroup from 2015-2024. Bars represent observed data by age in years, colored lines represent the estimated smooth incidence rate by age depending on the period (pre-/post- and pandemic).*

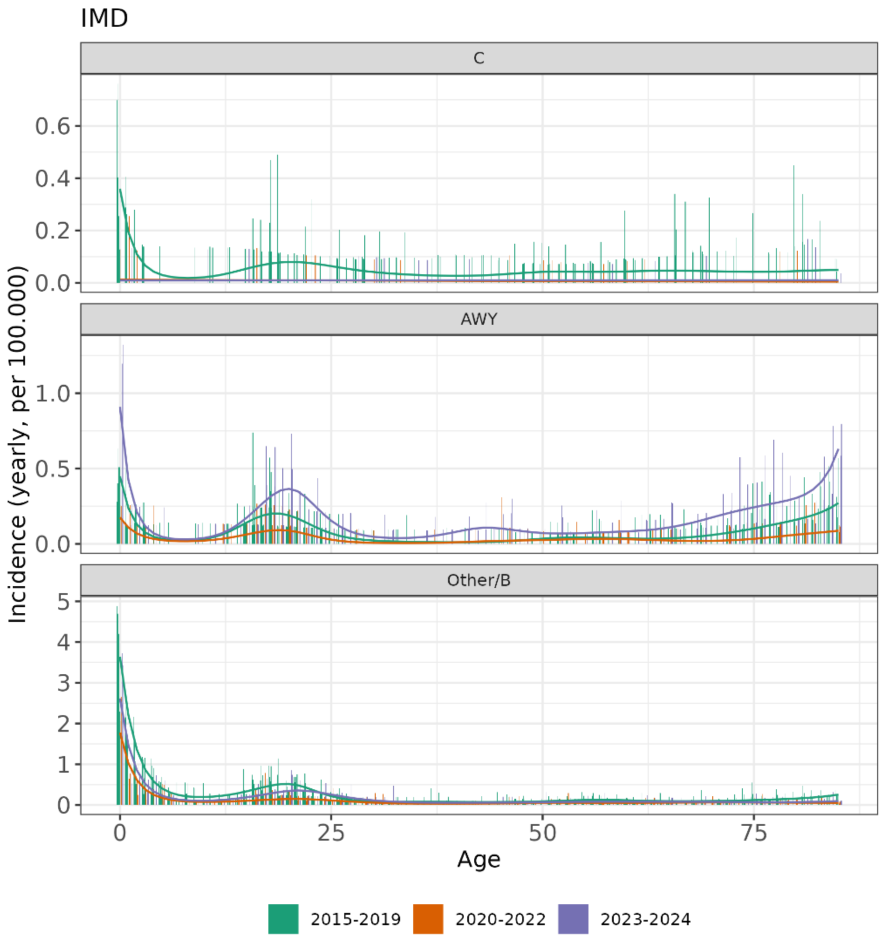


*Supplementary table M2: Vaccination parameters in the static model.*

| Strategy i | $vacc\_age_i$ | $upt_i$ | $ve_i^{MenC}$ | $ve_i^{MenAWY}$ | $r_i$ |
|---|---|---|---|---|---|
| 1) MenC infant | 1.5 | 0.80 | 1 | 0 | 1/4 |
| 2) No MenC/MenACWY | - | - | - | - | - |
| 3) MenACWY infant | 1.5 | 0.80 | 1 | 1 | 1/4 |
| 4) MenACWY 16y + infant[1] | $p_4^{MenX}(a) = p_3^{MenX}(a) + p_5^{MenX}(a)$ | | | | |
| 5) MenACWY 16y | 16 | 0.40 | 1 | 1 | 1/10 |

[1]The age-specific protection for strategy 4 is not calculated separately but derived from adding the age-specific protection of strategies 3 and 5.

### Supplementary material: Results

#### Epidemiological trends

*Supplementary figure 1: IMD incidence in Germany by age and serogroup in different time periods (columns) from 2002-2024. Bars represent the IMD incidence per 100,000 person-years in 1-year age-groups. First row shows incidence aggregated over all serogroups, and rows 2-4 for serogroups MenC, MenAWY and Other/B. Note the different y-axis range in the rows.*

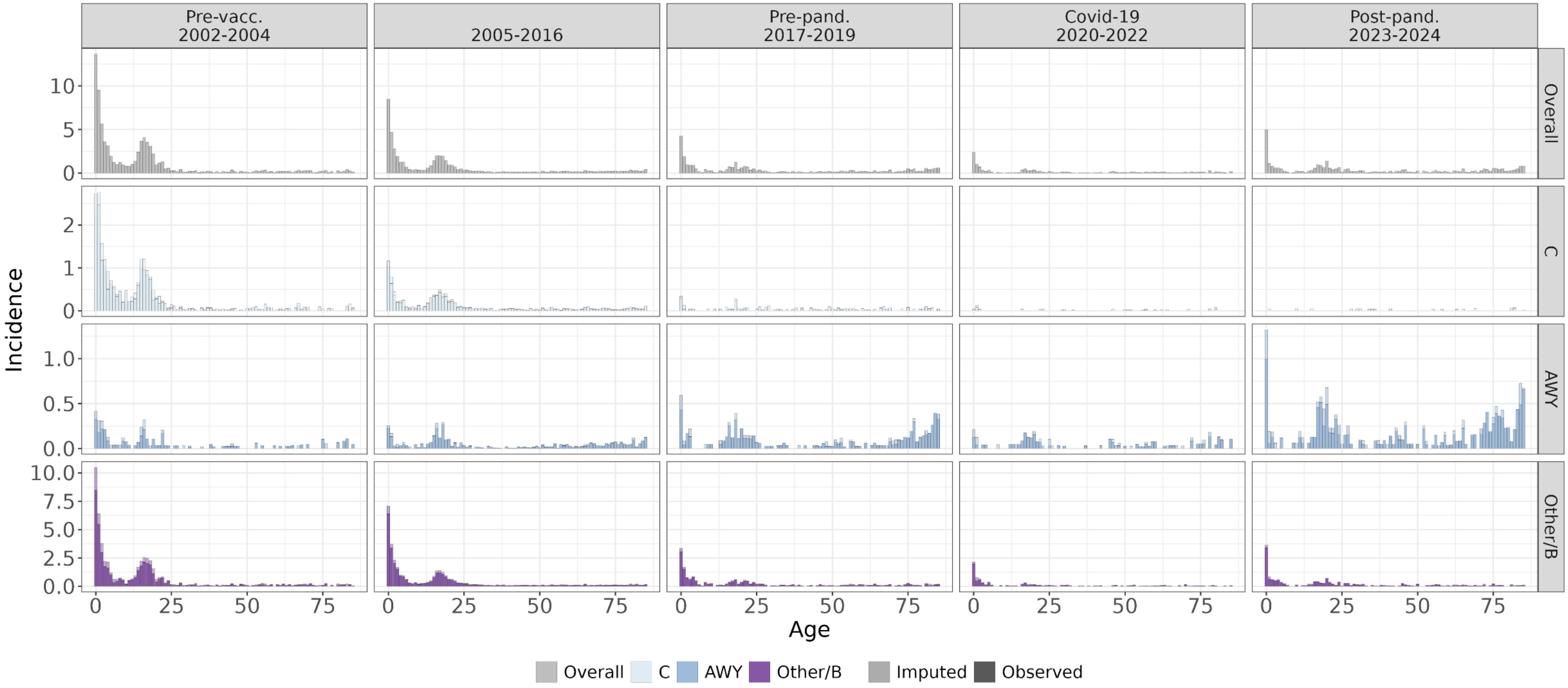

*Supplementary figure 2: IMD incidence in Germany by year and serogroup in 5 age-groups (0, 1-5, 6-15. 16-25, and 26+) from 2002-2024. Bars represent the yearly IMD incidence per 100,000 individuals. Color-coded are cases with reported and imputed serogroup, respectively. Numbers correspond to absolute case counts. (reported + imputed serogroup) Dotted lines mark period of SARS-CoV-2 pandemic (2020-2022). Note different y-axis range for Other/B serogroup compared to MenC and MenAWY.*

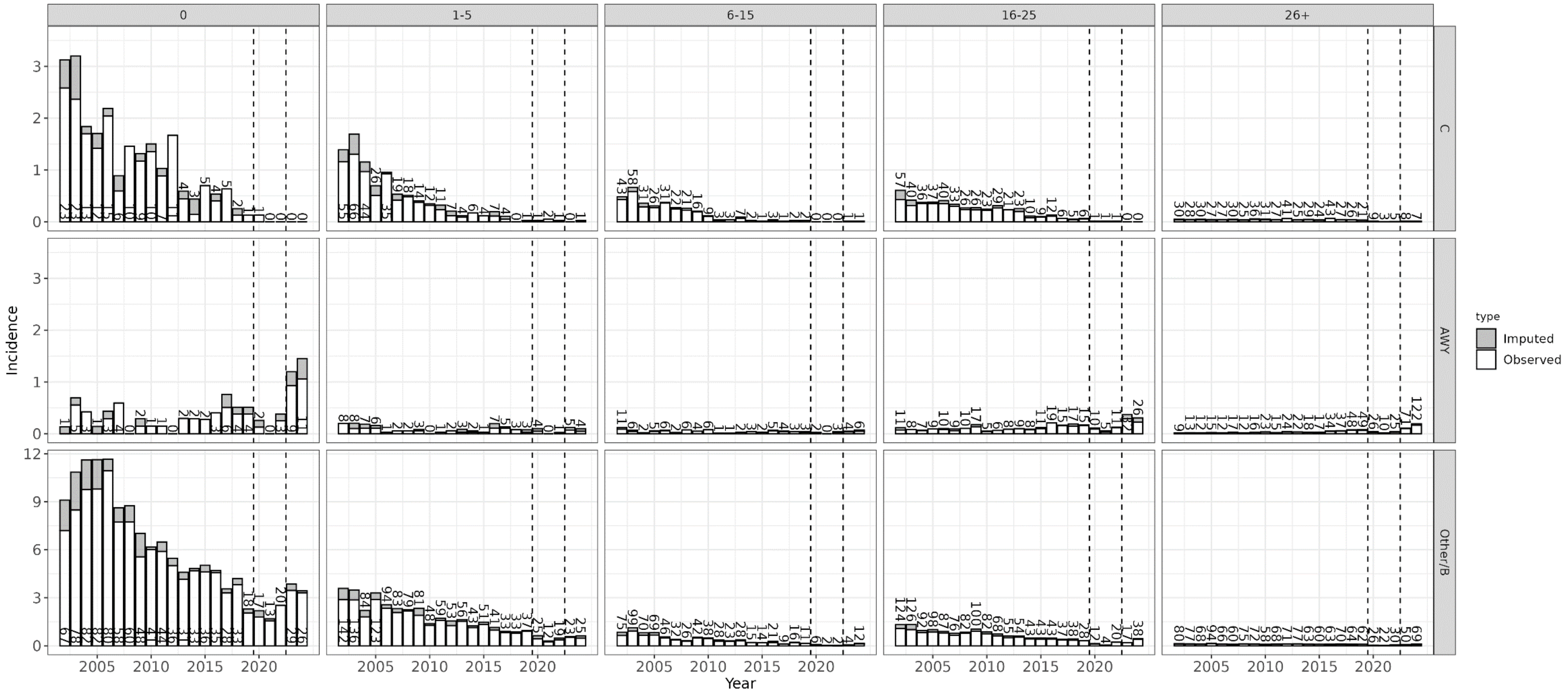


Children aged 1-5 (column 2) were the direct target group of the MenC toddler vaccination recommendation implemented in 2006. MenC incidence decreased in this group (as well as the other age groups) continuously since then. Starting from 2016, MenAWY case numbers were on a similar or higher level in in children aged 1-5 compared to MenC. Post-pandemic, MenAWY was more frequent among children 1-5 years as compared to MenC (with 5 observed and 4 imputed MenAWY cases and 1 MenC case for the years 2023 and 2024). Post-pandemic MenACWY incidence was higher in adolescents than in toddlers.

### Simulation of vaccination strategies

*Supplementary table 1: Yearly expected number of prevented IMD cases, vaccination numbers, and the NNV to prevent one IMD cases for 4 vaccination strategies compared to "no vaccination". Results are given for all serogroups and excluding serogroup "Other/B" (i.e., assuming no serogroup replacement). Sub-table A shows results for a 10-year simulation period, Sub-table B for a 30-year simulation.*

| A) 10-year simulation | | | | |
|---|---|---|---|---|
| Strategy | Serogroup | Prev. IMD | Vacc. Number | NNV |
| MenC infant | All sero. | 2.9 (2.2, 3.7) | 618,000 | 210,000 (170,000, 280,000) |
| | w\o Other/B | 2.9 (2.2, 3.7) | 618,000 | 210,000 (170,000, 280,000) |
| MenACWY infant | All sero. | 5.3 (4.4, 6.3) | 618,000 | 120,000 (98,000, 140,000) |
| | w\o Other/B | 5.3 (4.4, 6.3) | 618,000 | 120,000 (98,000, 140,000) |
| MenACWY infant + 16y | All sero. | 9.7 (8.5, 11.3) | 951,000 | 98,000 (84,000, 110,000) |
| | w\o Other/B | 11.7 (10.5, 13.4) | 951,000 | 81,000 (71,000, 91,000) |
| MenACWY 16y | All sero. | 4.6 (3.9, 5.4) | 334,000 | 72,000 (61,000, 86,000) |
| | w\o Other/B | 6.5 (5.7, 7.5) | 334,000 | 51,000 (45,000, 59,000) |
| **B) 30-year simulation** | | | | |
| Strategy | Serogroup | Prev. IMD | Vacc. Number | NNV |
| MenC infant | All sero. | 2.6 (1.8, 3.8) | 582,000 | 220,000 (150,000, 330,000) |
| | w\o Other/B | 2.7 (1.8, 3.9) | 582,000 | 220,000 (150,000, 330,000) |
| MenACWY infant | All sero. | 6.1 (5.1, 7.2) | 582,000 | 96,000 (81,000, 110,000) |
| | w\o Other/B | 6.4 (5.4, 7.5) | 582,000 | 91,000 (77,000, 110,000) |
| MenACWY infant + 16y | All sero. | 3.8 (-0.9, 7.6) | 911,000 | 240,000 (120,000, -*) |
| | w\o Other/B | 22.2 (19.2, 25) | 911,000 | 41,000 (36,000, 47,000) |
| MenACWY 16y | All sero. | 0.1 (-4, 3.2) | 328,000 | 3,700,000 (100,000, -*) |
| | w\o Other/B | 17.1 (14.8, 19.9) | 328,000 | 19,000 (16,000, 22,000) |

* NNV undefined in case of more IMD cases in simulation scenario compared to reference scenario

*Supplementary figure 3: Simulated IMD in Germany over 30-year simulation period. Panel A shows expected IMD counts for simulations of 4 different vaccination strategies (color-coded) in 3 sero-groups (rows), panel B shows an additional separation into 5 age-groups (columns). Panel C shows yearly changes in expected IMD counts by sero- and panel D by sero- and age-group compared to a "no vaccination" scenario for the four strategies.*

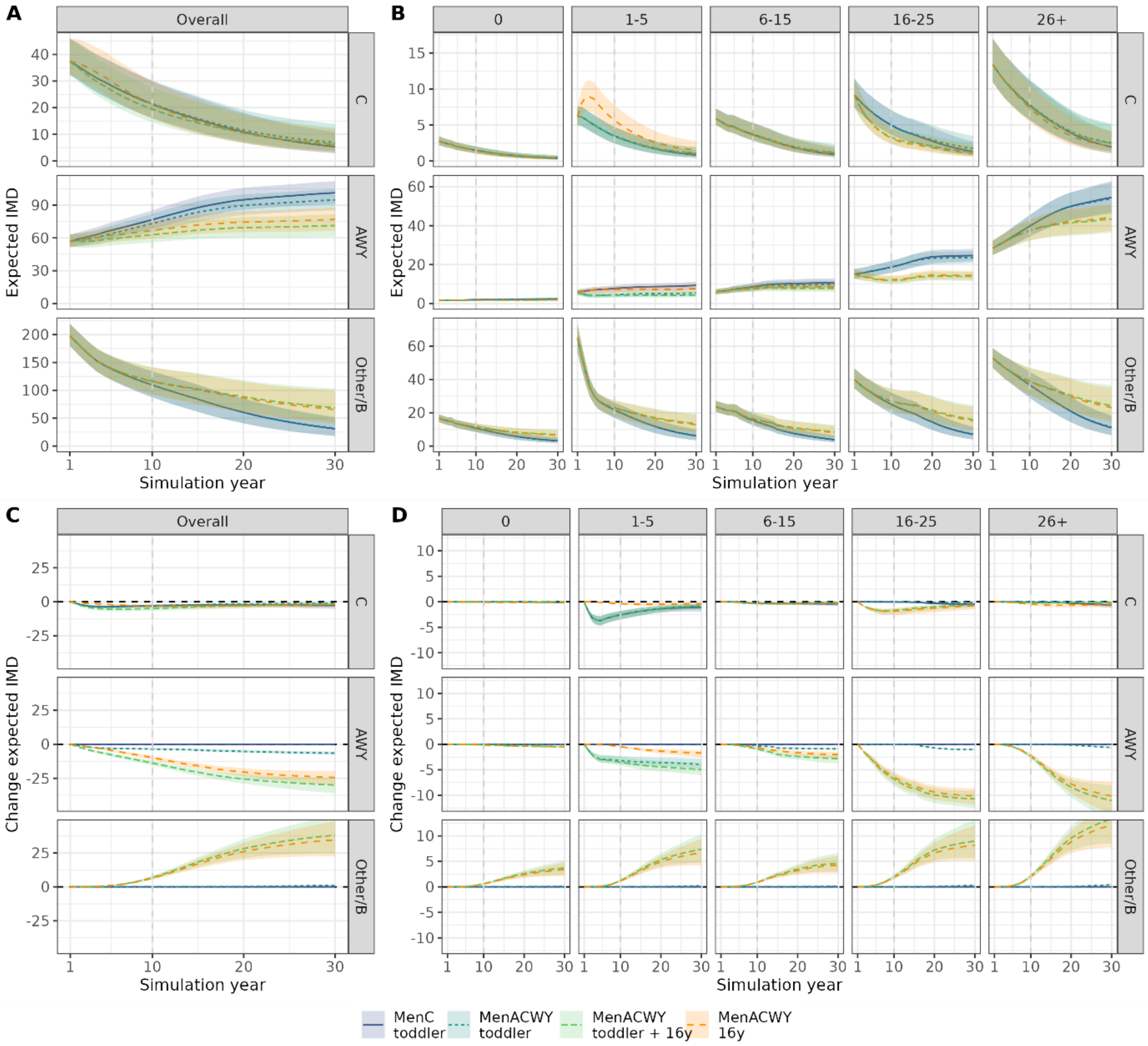

*Supplementary figure 4: Prevented number of IMD cases and NNV for primary MenACWY adolescent vaccination at different age of vaccination and different levels of uptake in Germany based on the static model.*

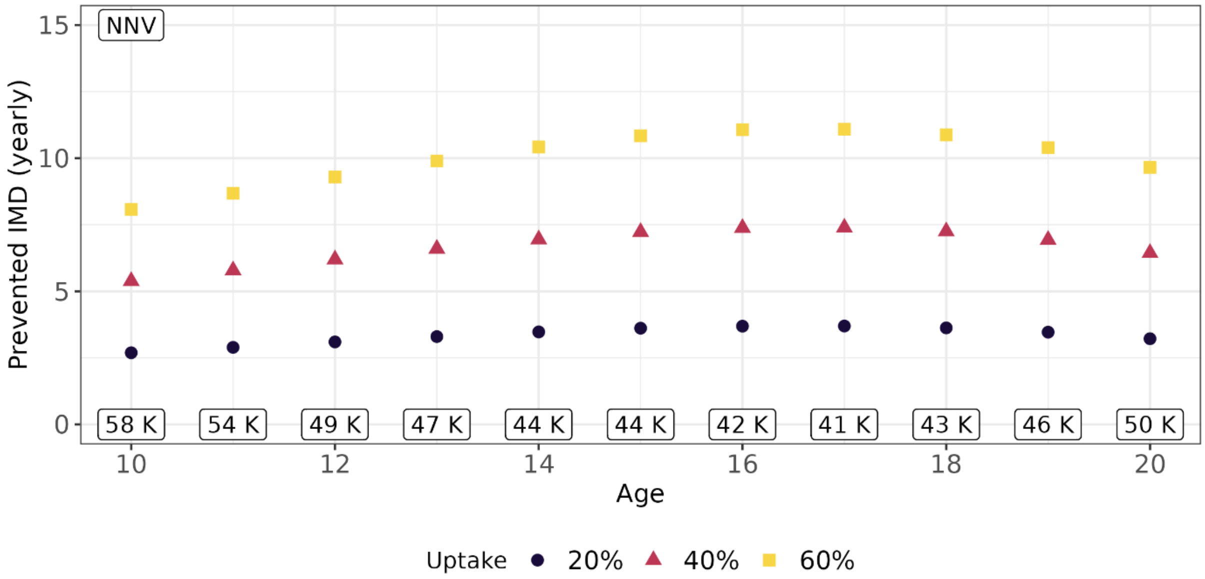


K: kilo (i.e., in thousands). Vaccination at age 17 was most efficient with a NNV of 41.000 vaccinations to prevent one IMD case. Vaccination at a younger age reduces efficiency, the number of vaccinations to prevent on IMD case was, e.g., at age 12 years 17% higher compared to the scenario of vaccination at 16 years (as in the main analysis). However, the overall reduction in the disease burden could be enhanced if a higher vaccination uptake is achieved when vaccination is given at a younger age.
